# Knowledge of novel coronavirus (SARS-COV-2) among a Georgian population

**DOI:** 10.1101/2020.05.14.20101642

**Authors:** Maia Butsashvili, Lasha Gulbiani, George Kanchelashvili, Marika Kochlamazashvili, George Nioradze, George kamkamidze

**Author notes:** **Corresponding author:** Maia Butsashvili, **Corresponding author email:**.

## Abstract

**Introduction:** Georgia confirmed its first case of SARS-COV-2 infection on February 26, 2020. Despite the government’s proactive measures during the early stages of the epidemic, number of new infections of SARS-COV-2 is increasing and by March 31, a total of 110 cases have been reported. Limited understanding about epidemics can lead to panic and disrupt public health efforts to contain transmission. Thus, it is very important to understand the perceptions of the population regarding the disease and perceived level of government preparedness to fight against the spread of infection. This study reports results of a survey designed to understand attitudes and knowledge regarding SARS-COV-2 virus among Georgian population, including health care workers (HCWs).

**Materials and methods:** The online survey was conducted using a Facebook advertisement. The target was the whole country and the language used was Georgian. We collected information on demographic data, knowledge of symptoms and transmission modes of coronavirus, perceived differences between coronavirus and influenza, availability of antiviral medication and vaccination. We also included questions to capture the Georgian population’s perceptions about government preparedness to combat the new coronavirus.

**Results:** The survey was open for three days (March 2–4, 2020). 5228 participants completed the survey. Of these, 40.3% were 25–45 years old and 58.2% were female. 20.7% of respondents had university degree and 10.3% were HCWs. For 25.8% of respondents, COVID-19 and influenza are the same diseases; 10.9% did not know if they are different. The majority correctly identified the transmission route and symptoms (96.9% and 98.0%, respectively). Regarding physical distancing, 13.2% indicated they would attend public events if needed even if they had COVID-19 symptoms. 19.1% think that Georgia is ready for COVID 19 epidemic, while according to 55% the county is not ready, but HCWs are trying hard to respond to this challenge properly. For 18% response is inadequate. There was no difference in knowledge between HCWs, non-HCWs and unemployed. 20% of HCWs as well as other study subjects believe that SARS-COV-2 vaccine and medications do exist but are simply not available in Georgia.

**Conclusion:** One in five Georgians believe that there is a vaccine and medication to treat coronavirus, but that it is not available in the country. Given that information regarding coronavirus is changing very rapidly, the need to reach people with time-sensitive educational messages as well as prevention strategies is vital.

## Introduction

Three months have elapsed since discovery of the novel coronavirus causing severe acute respiratory syndrome and classified as SARS-COV-2 [1–3].

Georgia, an Eastern European country, confirmed its first case of SARS-COV-2 infection on February 26, 2020. Despite the government’s proactive measures during the early stages of the epidemic, such as restriction of air travel, the number of new infections of SARS-COV-2 is increasing and by March 31, a total of 110 cases have been reported [4].

Limited understanding about epidemics can lead to panic and disrupt public health efforts to contain transmission as people crowd into grocery stores and use public transportation to prepare for an undefined period of lockdown [5]. Thus, it is very important to understand the perceptions of the population regarding the disease and pandemic, and the perceived level of government preparedness to fight against the spread of infection. This study reports results of a survey designed to understand attitudes and knowledge regarding SARS-COV-2 virus and perceptions of preventive measures among the Georgian population, including health care workers (HCWs).

## Materials and methods

The online survey was conducted using a Facebook advertisement, which included the title, body text, the banner and the link to the questionnaire. The target was the whole country and the language used was Georgian. The study was approved by the Institutional Review Board of Health Research Union.

We collected information on demographic data (age, gender, marital status, education, employment status), knowledge of symptoms and transmission modes of coronavirus, perceived differences between coronavirus and influenza, availability of antiviral medication and vaccination. We also included questions to capture the Georgian population’s perceptions about government preparedness to combat the new coronavirus.

## Results

The survey was open for three days (March 2–4, 2020), during which time 5228 participants completed the survey. Of these, 40.3% (n=2106) were 25–45 years old, 58.2% (n=3042) were female, and 46.7% (n=2440) were married. One in five (20.7%, n=1080) of respondents had a university degree (Masters or Doctoral) and 10.3% (n=536) were working in the health care field (HCWs).

For 25.8% (n=1348) of respondents, a perception exists that COVID-19 and influenza are the same diseases; an additional 10.9% (n=568) did not know if they are different. In response to the question “Are you afraid of getting infected with SARS-COV-2?” almost half of study participants (46.3%) said “no.” The majority of survey respondents correctly identified the transmission route and symptoms of the new coronavirus (96.9% and 98.0%, respectively). Respondents had little knowledge regarding antiviral medications and vaccines against COVID-19. Specifically, 41.4% (n=2164) did not correctly respond about the existence of antivirals and only 65.6% (n=3429) were aware that a vaccine does not exist. Nearly half (45.3%) of the respondents reported that COVID-19 mortality rates vary from 2 to 5% (Table 1).

**Table 1.** Knowledge regarding COVID-19

| Characteristic | N | % |
| --- | --- | --- |
| **Are coronavirus and influenza the same diseases?** |  |  |
| Yes | 1348 | 25.8 |
| No | 3312 | 63.4 |
| Don't know | 568 | 10.9 |
| **Are you afraid of getting infected with SARS-COV-2?** |  |  |
| Yes | 2288 | 43.8 |
| No | 2418 | 46.3 |
| Don't know | 522 | 10.0 |
| **Is your fear caused by information in the media and social network regarding COVID-19?** |  |  |
| Yes | 1050 | 45.9 |
| No, my fear is not relevant to these sources | 1120 | 49.0 |
| I don't know | 118 | 5.2 |
| **SARS-COV-2 transmission route is:** |  |  |
| Droplets | 5064 | 96.9 |
| Sexual intercourse | 44 | 0.8 |
| Blood borne | 10 | 0.2 |
| I don't know | 110 | 2.1 |
| **COVID-19 symptoms are:** |  |  |
| Dizziness, loss of appetite, losing weight | 14 | 0.3 |
| Bloody cough, frequent urination, enlarged lymph nodes | 10 | 0.2 |
| Fever, cough, shortness of breath | 5126 | 98.0 |
| I don't know | 78 | 1.5 |
| **Does a vaccine against SARS-COV-2 exist?** |  |  |
| Yes, and it is available in Georgia | 140 | 2.7 |
| Yes, but it isn't available in Georgia | 858 | 16.4 |
| No | 3429 | 65.6 |
| I don't know | 800 | 15.3 |
| **Do antiviral medications against SARS-COV-2 exist?** |  |  |
| Yes | 880 | 16.8 |
| No | 3063 | 58.6 |
| Don't know | 1284 | 24.6 |
| **COVID-19 related mortality is:** |  |  |
| 0 – 1.5% | 1100 | 21.0 |
| 2 – 5% | 2368 | 45.3 |
| >5% | 682 | 13.0 |
| I don't know | 1078 | 20.6 |

The majority of participants (95.2%) reported that they would visit a medical facility if symptoms presented. Regarding precautions, 58.3% (n=3044) reported using masks in public places. A large proportion of surveyed individuals (75.4%) declared that wearing a mask is partially protective against SARS-COV-2. Regarding preparations, we asked “have you stocked up on food for current situation related to coronavirus epidemic?” Over half of respondents (53.6%, n=2800) reported that they did not think it was necessary. Regarding physical distancing strategies to reduce transmission, 13.2% (n=688) indicated they would attend public events if needed even if they had COVID-19 symptoms. 19.1% (n=996) of study participants think that Georgia is ready for COVID 19 epidemic, while according to 55% (n=2898) the county is not ready, but health care institutions are trying hard to respond to this challenge properly. For 18% (n=954) of study subjects, response to the current epidemic in the country is inadequate. Very few (1.1%, n=56) of study subjects visited countries with a high prevalence of SARS-COV-2 during the last month (February 2020) (Table 2).

**Table 2.**
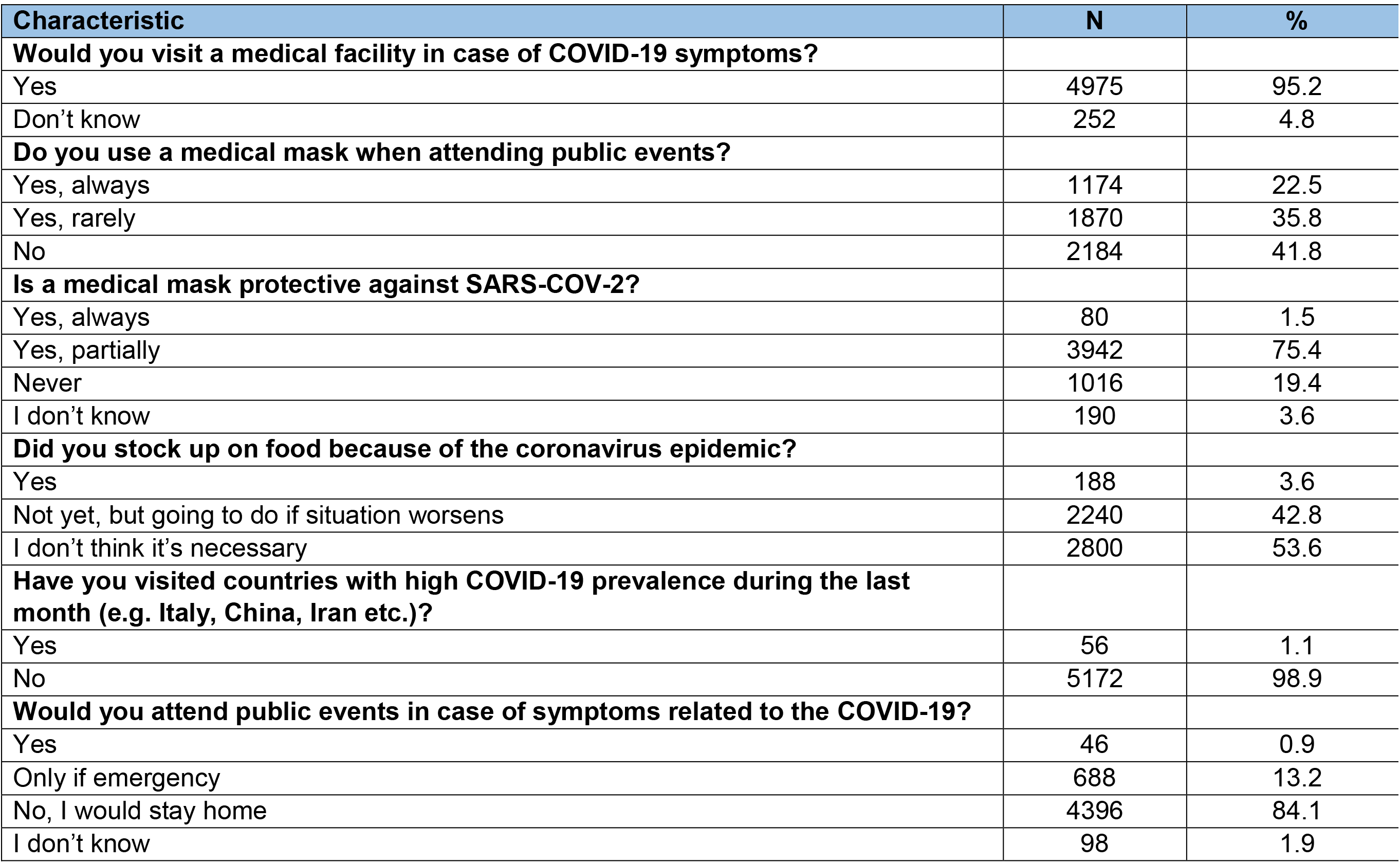
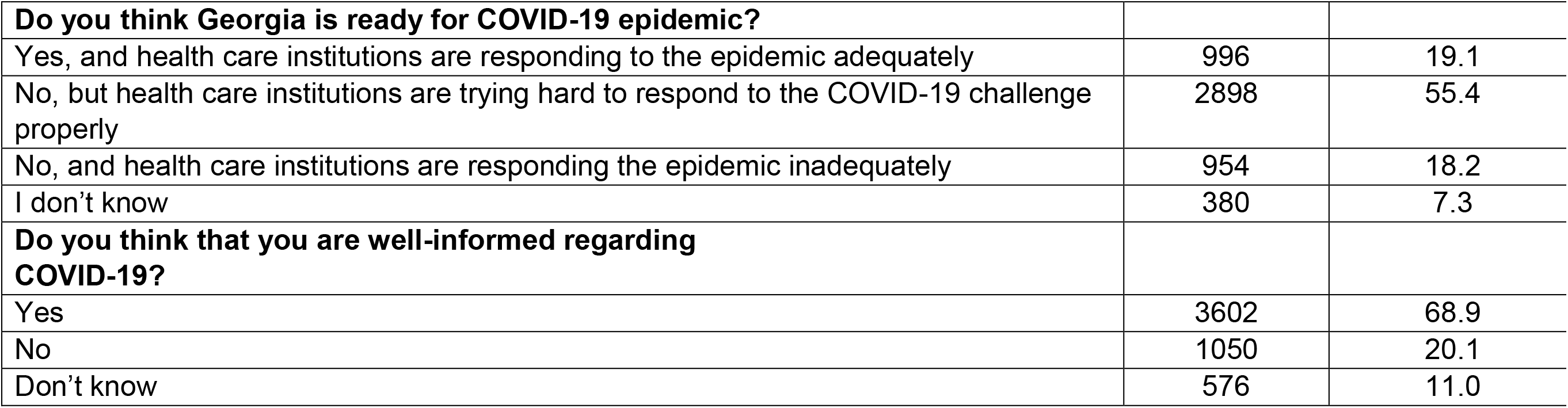
Attitude and perception towards COVID-19.

| Characteristic | N | % |
| --- | --- | --- |
| **Would you visit a medical facility in case of COVID-19 symptoms?** |  |  |
| Yes | 4975 | 95.2 |
| Don't know | 252 | 4.8 |
| **Do you use a medical mask when attending public events?** |  |  |
| Yes, always | 1174 | 22.5 |
| Yes, rarely | 1870 | 35.8 |
| No | 2184 | 41.8 |
| **Is a medical mask protective against SARS-COV-2?** |  |  |
| Yes, always | 80 | 1.5 |
| Yes, partially | 3942 | 75.4 |
| Never | 1016 | 19.4 |
| I don't know | 190 | 3.6 |
| **Did you stock up on food because of the coronavirus epidemic?** |  |  |
| Yes | 188 | 3.6 |
| Not yet, but going to do if situation worsens | 2240 | 42.8 |
| I don't think it's necessary | 2800 | 53.6 |
| **Have you visited countries with high COVID-19 prevalence during the last month (e.g. Italy, China, Iran etc.)?** |  |  |
| Yes | 56 | 1.1 |
| No | 5172 | 98.9 |
| **Would you attend public events in case of symptoms related to the COVID-19?** |  |  |
| Yes | 46 | 0.9 |
| Only if emergency | 688 | 13.2 |
| No, I would stay home | 4396 | 84.1 |
| I don't know | 98 | 1.9 |
| **Do you think Georgia is ready for COVID-19 epidemic?** |  |  |
| Yes, and health care institutions are responding to the epidemic adequately | 996 | 19.1 |
| No, but health care institutions are trying hard to respond to the COVID-19 challenge properly | 2898 | 55.4 |
| No, and health care institutions are responding the epidemic inadequately | 954 | 18.2 |
| I don't know | 380 | 7.3 |
| **Do you think that you are well-informed regarding COVID-19?** |  |  |
| Yes | 3602 | 68.9 |
| No | 1050 | 20.1 |
| Don't know | 576 | 11.0 |

Younger respondents differentiated COVID-19 from influenza better than their older counterparts. Among those aged 16–24 years, 67.1% (n=1438) indicated that COVID-19 and influenza are not the same, followed by 62.4% (n=1314) of participants aged 25 to 45 and 57.1% (n=560) of those older than 45 years (*p*<0.0001) (Table 3).

**Table 3.** Knowledge, attitudes and perceptions regarding COVID-19 by age.

| Characteristic | Total |  | 16-24 |  | 25-45 |  | ≥46 |  | *p*<br>value |
| --- | --- | --- | --- | --- | --- | --- | --- | --- | --- |
|  | N | % | N | % | N | % | N | % |  |
| **Are coronavirus and influenza the same diseases?** |  |  |  |  |  |  |  |  |  |
| Yes | 1348 | 25.8 | 454 | 21.2 | 556 | 26.4 | 338 | 34.5 | <0.001 |
| No | 3312 | 63.4 | 1438 | 67.1 | 1314 | 62.4 | 560 | 57.1 |  |
| Don't know | 568 | 10.9 | 250 | 11.7 | 236 | 11.2 | 82 | 8.4 |  |
| **Are you afraid of getting infected with SARS-COV-2?** |  |  |  |  |  |  |  |  |  |
| Yes | 2288 | 43.8 | 876 | 40.9 | 966 | 45.9 | 446 | 45.5 | <0.001 |
| No | 2418 | 46.3 | 1058 | 49.4 | 944 | 44.8 | 416 | 42.4 |  |
| Don't know | 522 | 10.0 | 208 | 9.7 | 196 | 9.3 | 118 | 12.0 |  |
| **Is your fear caused by the media and social network regarding COVID-19?** |  |  |  |  |  |  |  |  |  |
| Yes | 1050 | 45.9 | 424 | 48.4 | 472 | 48.9 | 154 | 34.5 | <0.001 |
| No, my fear is relevant | 1120 | 49.0 | 404 | 46.1 | 436 | 45.1 | 280 | 62.8 |  |
| I don't know | 118 | 5.2 | 48 | 5.5 | 58 | 6.0 | 12 | 2.7 |  |
| **SARS-COV-2 transmission route is:** |  |  |  |  |  |  |  |  |  |
| Droplets | 5064 | 96.6 | 2052 | 95.8 | 2042 | 97.0 | 970 | 99.0 | <0.001 |
| Sexual intercourse | 44 | 0.8 | 30 | 1.4 | 12 | 0.6 | 2 | 0.2 |  |
| Blood Bourn | 10 | 0.2 | 6 | 0.3 | 2 | 0.1 | 2 | 0.2 |  |
| I don't know | 110 | 2.1 | 54 | 2.5 | 50 | 2.4 | 6 | 0.6 |  |
| **COVID-19 symptoms are:** |  |  |  |  |  |  |  |  |  |
| Dizziness, loss of appetite, losing weight | 14 | 0.3 | 14 | 0.7 | 0 | 0.0 | 0 | 0.0 | <0.001 |
| Bloody cough, frequent urination, lymph nodes | 10 | 0.2 | 8 | 0.4 | 0 | 0.0 | 2 | 0.2 |  |
| Fever, cough, shortness of breath | 5126 | 98.0 | 2082 | 97.2 | 2074 | 98.5 | 970 | 99.0 |  |
| I don't know | 78 | 1.5 | 38 | 1.8 | 32 | 1.5 | 8 | 0.8 |  |
| **Does a vaccine against SARS-COV-2 exist?** |  |  |  |  |  |  |  |  |  |
| Yes, and it is available in Georgia | 140 | 2.7 | 90 | 4.2 | 42 | 2.0 | 8 | 0.8 | <0.001 |
| Yes, but isn't available in Georgia | 858 | 16.4 | 516 | 24.1 | 272 | 12.9 | 70 | 7.1 |  |
| No | 3430 | 65.6 | 1134 | 52.9 | 1506 | 71.5 | 790 | 80.6 |  |
| I don't know | 800 | 15.3 | 402 | 18.8 | 286 | 13.6 | 112 | 11.4 |  |
| **Do antiviral medications against SARS-COV-2 exist?** |  |  |  |  |  |  |  |  |  |
| Yes | 880 | 16.8 | 438 | 20.4 | 326 | 15.5 | 116 | 11.8 | <0.001 |
| No | 3064 | 58.6 | 1068 | 49.9 | 1312 | 62.3 | 684 | 69.8 |  |
| Don't know | 1284 | 24.6 | 636 | 29.7 | 468 | 22.2 | 180 | 18.4 |  |
| **Would you visit a medical facility for diagnosis if you suspect you have COVID-19?** |  |  |  |  |  |  |  |  |  |
| Yes | 4976 | 95.2 | 2030 | 94.8 | 2006 | 95.3 | 940 | 95.9 | 0.374 |
| No | 0 | 0.0 | 0 | 0.0 | 0 | 0.0 | 0 | 0.0 |  |
| Don't know | 252 | 4.8 | 112 | 5.2 | 100 | 4.7 | 40 | 4.1 |  |
| **COVID-19-related mortality is:** |  |  |  |  |  |  |  |  |  |
| 0 – 1.5% | 1100 | 21.0 | 382 | 17.8 | 464 | 22.0 | 254 | 25.9 | <0.001 |
| 2 – 5% | 2368 | 45.3 | 888 | 41.5 | 988 | 46.9 | 492 | 50.2 |  |
| >5% | 682 | 13.0 | 358 | 16.7 | 212 | 10.1 | 112 | 11.4 |  |
| I don't know | 1078 | 20.6 | 514 | 24.0 | 442 | 21.0 | 122 | 12.4 |  |
| **Do you use a medical mask when attending public events?** |  |  |  |  |  |  |  |  |  |
| Yes, always | 1174 | 22.5 | 534 | 24.9 | 468 | 22.2 | 172 | 17.6 | <0.001 |
| Yes, rarely | 1870 | 35.8 | 800 | 37.3 | 728 | 34.6 | 342 | 34.9 |  |
| No | 2184 | 41.8 | 808 | 37.7 | 910 | 43.2 | 466 | 47.6 |  |
| **Is a medical mask protecting against SARS-COV-2?** |  |  |  |  |  |  |  |  |  |
| Yes, always | 80 | 1.5 | 28 | 1.3 | 32 | 1.5 | 20 | 2.0 | 0.066 |
| Yes, partially | 3942 | 75.4 | 1648 | 76.9 | 1550 | 73.6 | 744 | 75.9 |  |
| Never | 1016 | 19.4 | 394 | 18.4 | 448 | 21.3 | 174 | 17.8 |  |
| I don't know | 190 | 3.6 | 72 | 3.4 | 76 | 3.6 | 42 | 4.3 |  |
| **Did you stock up on food because of coronavirus epidemic?** |  |  |  |  |  |  |  |  |  |
| Yes | 188 | 3.6 | 70 | 3.3 | 78 | 3.7 | 40 | 4.1 | 0.216 |
| Not yet, but going to do if situation worsens | 2240 | 42.8 | 900 | 42.0 | 894 | 42.5 | 446 | 45.5 |  |
| I don't think it's necessary | 2800 | 53.6 | 1172 | 54.7 | 1134 | 53.8 | 494 | 50.4 |  |
| **Do you think Georgia is ready for the COVID-19 epidemic?** |  |  |  |  |  |  |  |  |  |
| Yes, and health care institutions are responding the epidemic adequately | 996 | 19.1 | 314 | 14.7 | 416 | 19.8 | 266 | 27.1 | <0.001 |
| No, but health care institutions are trying hard to response COVID-19 challenge properly | 2898 | 55.4 | 1092 | 51.0 | 1220 | 57.9 | 586 | 59.8 |  |
| No, and health care institutions are responding the epidemic inadequately | 954 | 18.2 | 578 | 27.0 | 308 | 14.6 | 68 | 6.9 |  |
| I don't know | 380 | 7.3 | 158 | 7.4 | 162 | 7.7 | 60 | 6.1 |  |
| **Would you attend public events if you have symptoms related to COVID-19?** |  |  |  |  |  |  |  |  |  |
| Yes | 46 | 0.9 | 24 | 1.1 | 16 | 0.8 | 6 | 0.6 | <0.001 |
| No, I would stay home | 4396 | 84.1 | 1710 | 79.8 | 1852 | 87.9 | 834 | 85.1 |  |
| Only if emergency | 688 | 13.2 | 358 | 16.7 | 210 | 10.0 | 120 | 12.2 |  |
| I don't know | 98 | 1.9 | 50 | 2.3 | 28 | 1.3 | 20 | 2.0 |  |
| **Have you visited countries with a high COVID-19 prevalence during last month (e.g. Italy, China, Iran etc.)?** |  |  |  |  |  |  |  |  |  |
| Yes | 56 | 1.1 | 32 | 1.5 | 14 | 0.7 | 10 | 1.0 | 0.31 |
| No | 5172 | 98.9 | 2110 | 98.5 | 2092 | 99.3 | 970 | 99.0 |  |

| Do you think that you are well-informed regarding COVID-19 epidemic? |  |  |  |  |  |  |  |  |  |
| --- | --- | --- | --- | --- | --- | --- | --- | --- | --- |
| Yes | 3602 | 68.9 | 1262 | 58.9 | 1550 | 73.6 | 790 | 80.6 | <0.001 |
| No | 1050 | 20.1 | 614 | 28.7 | 312 | 14.8 | 124 | 12.7 |  |
| Don't know | 576 | 11.0 | 266 | 12.4 | 244 | 11.6 | 66 | 6.7 |  |

We also found that educational attainment was positively associated with awareness regarding a vaccine and antiviral medications. The majority (82.2%, n=888) of respondents who had achieved a Masters or Doctoral degree correctly reported that a vaccine against SARS-COV-2 does not exist compared to 72.1% (n=1282) of those with a bachelor, 56.7% (n=632) of university students, and 50.0% (n=628) of high school graduates. Similarly, 70.6% (n=762) of postgraduates correctly responded that antiviral medications against SARS-COV-2 do not exist, followed by 62.1% (n=1104) of university graduates, 53.0% (n=590) of university students and 48.4% (n=608) of respondents with a high school education (*p*<0.0001).

Health care workers were less apprehensive about infection. In response to the question “Are you afraid of getting infected with COVID-19?” more than half (55.2%) of HCWs responded “no” compared to their unemployed (46.4%) or non-HCW counterparts (44.3%, *p*<0.0001). There was no detectable difference in knowledge about the virus or therapeutics between HCWs, non-HCWs and the unemployed. In fact, approximately 20% of HCWs as well as other study subjects believe that a SARS-COV-2 vaccine and medications do exist but are simply not available in Georgia (Table 4).

**Table 4.**
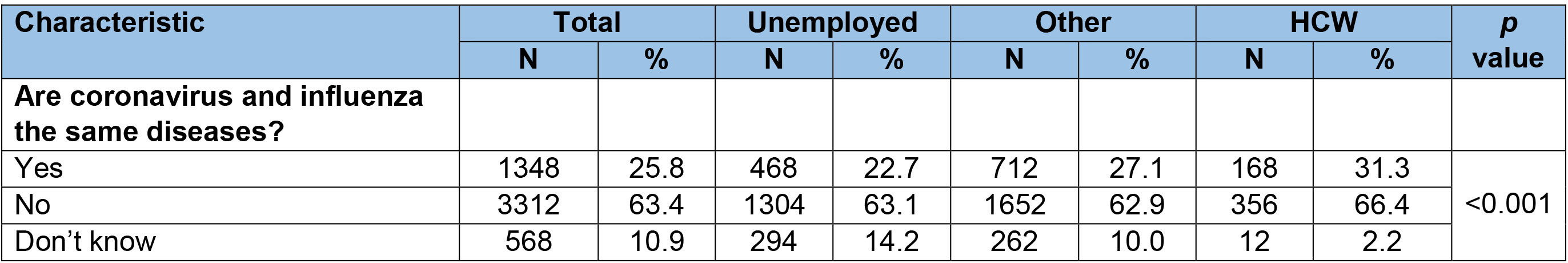
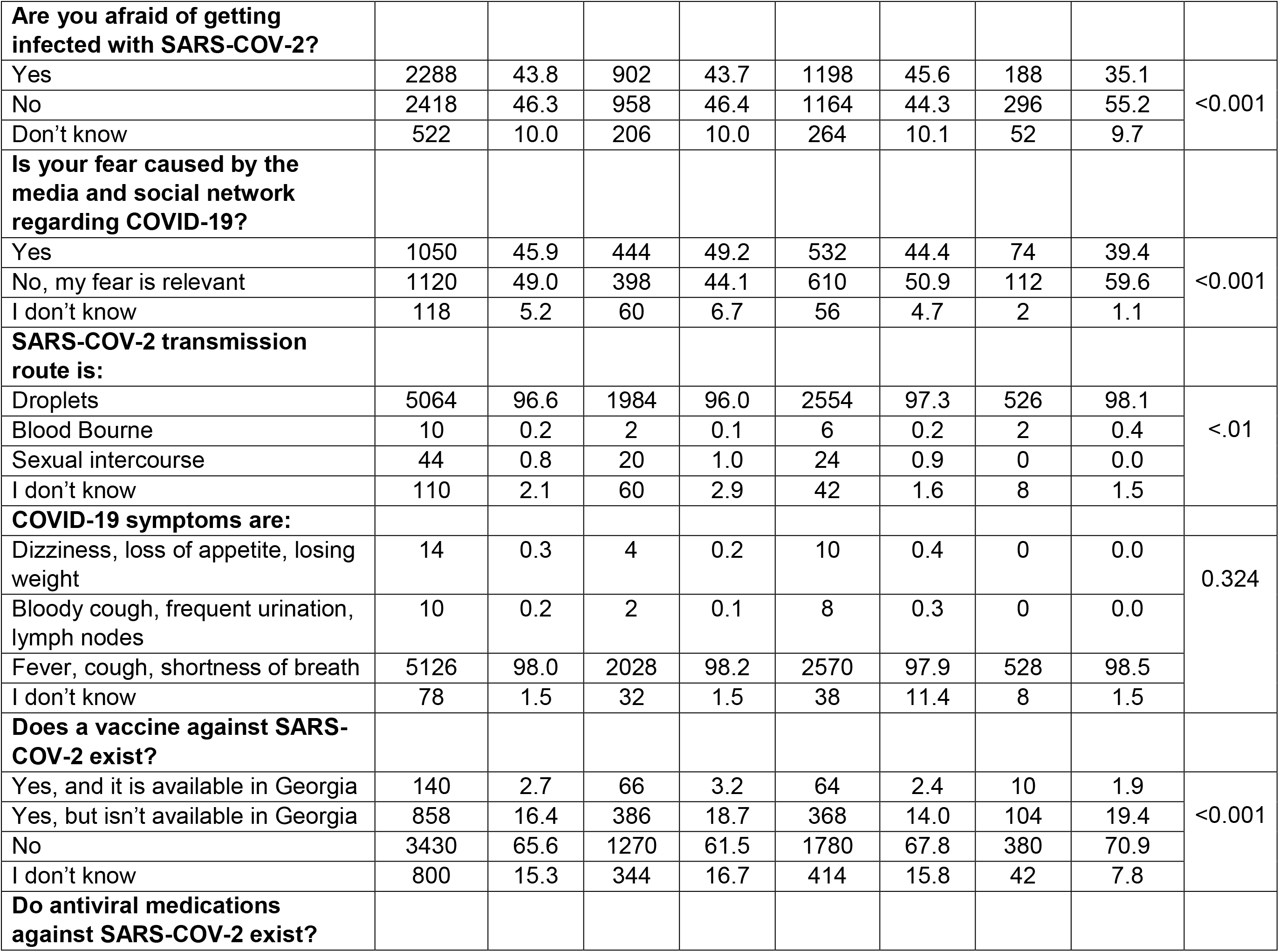
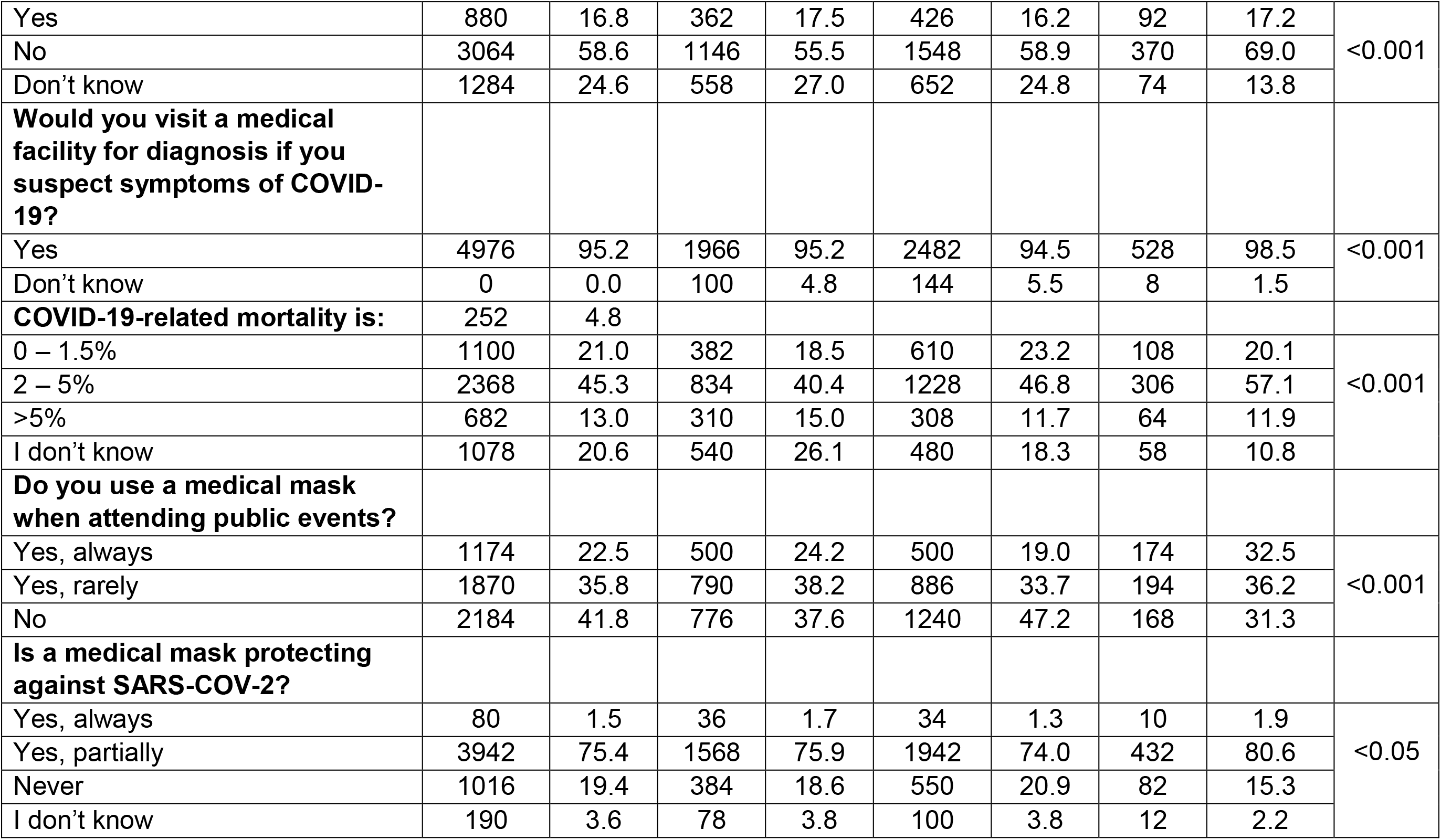
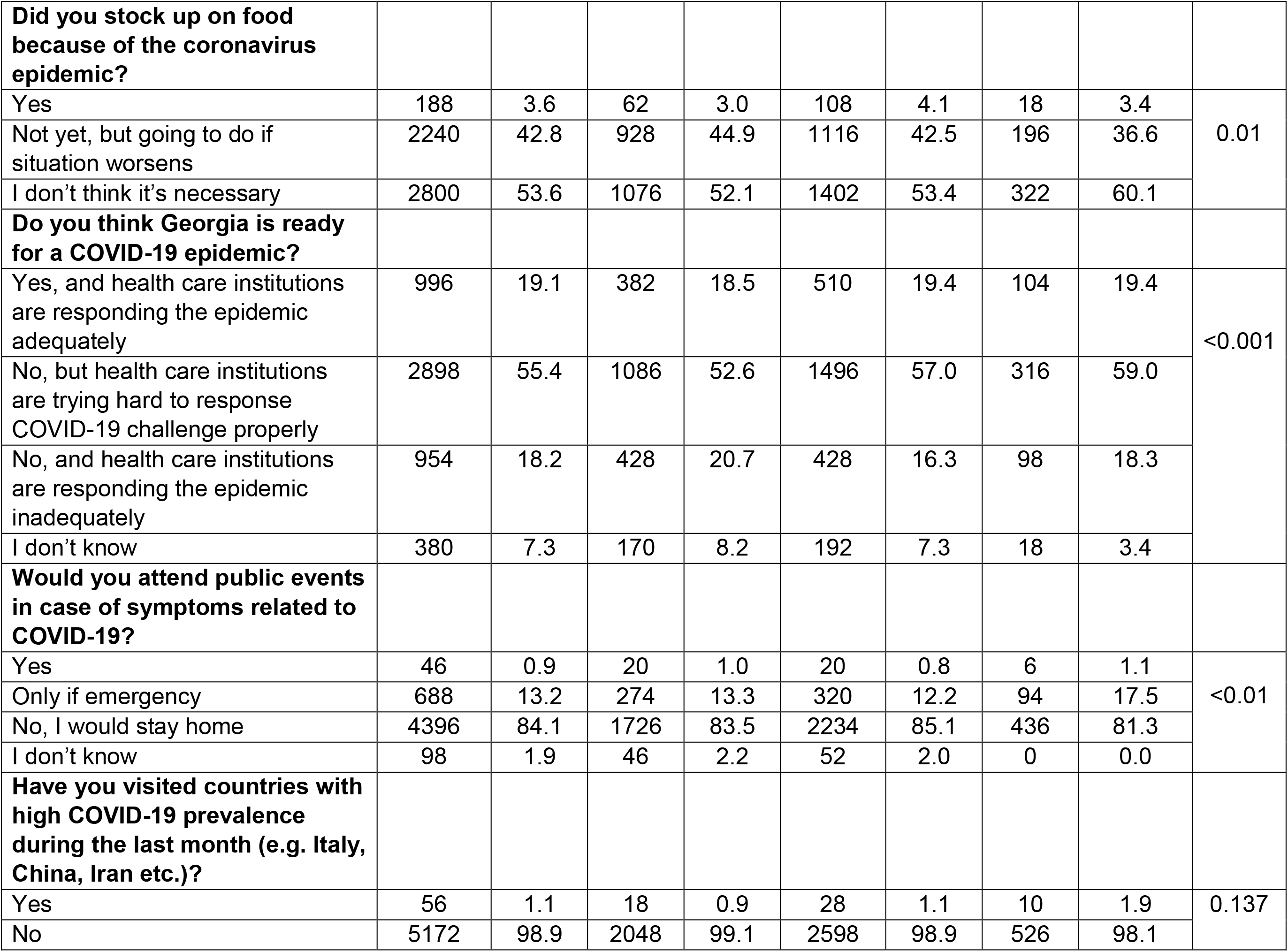
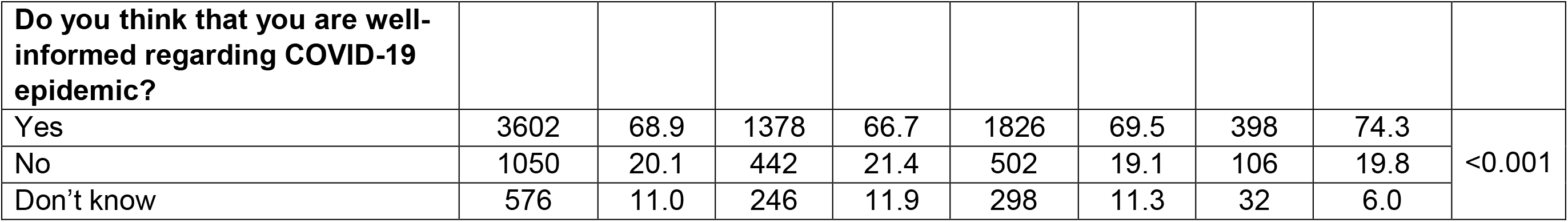
Knowledge, attitudes and perceptions regarding COVID-19 by profession.

## Discussion

We chose a convenience sample of Facebook participants for our survey because this method is quick, low-cost and reaches many participants [6–8]. During an epidemic, a face-to-face interview is not appropriate. Due to self-quarantine, telephone usage is increased, thus an online survey is most convenient to ensure all potential respondents have access to survey completion. The major limitation of this method is that internet access is differentially apportioned throughout the population. The Georgian Statistics Department estimated internet coverage in 2019 to be 79.3%, with 86% coverage in urban areas and 69.9% in rural areas [9]. We anticipate that Facebook accounts are more prevalent among the younger, educated population, as is reflected in our respondent demographics.

The level of knowledge was higher among older individuals, which is consistent with previous studies. An online survey about COVID-19 conducted in the United Kingdom during March 17–18 similarly demonstrated that older adults consider COVID-19 to be life-threatening [10]. However, overall awareness and appreciation of the risks appears to be higher among the UK respondents, 77% of whom worried about a coronavirus outbreak, compared to 44% of Georgian respondents.

According to our study, the media plays important role in disseminating information regarding the coronavirus pandemic, including among HCWs. This appears to be a similar trend found during previous outbreaks. For instance, in a study conducted during the SARS epidemic, 92% of participants in a KAP survey conducted in China reported that their primary source of information about the disease was television [11].

In conclusion, educational attainment and age are correlated with correct information about COVID-19. However, misinformation persists. One in five Georgians believe that there is a vaccine and medication to treat coronavirus, but that it is not available in the country. Training of HCWs is essential to improve their awareness level. More than 10% of Georgians would still attend a large public event, even with symptoms of COVID-19. The media is the primary source of information about COVID-19, widely relied upon by the general public as well as HCWs. Given that information regarding coronavirus is changing very rapidly, the need to reach people with time-sensitive educational messages as well as prevention strategies is vital.

## Data Availability

All data referred to in the manuscript is available upon request

